# Time-course responses following sports-related concussion: A multi-modality study

**DOI:** 10.1101/2025.02.02.25321562

**Authors:** Alan J Pearce, Kane Middleton, Anthea Clarke

## Abstract

**Objectives:** Using a repeated-measures design, we investigated reported symptoms, oculomotor, and neurophysiological responses for up to 26 days following a sports-related concussion.

**Methods:** Over the course of one season, 115 athletes (mean age 26.2 ± 5.4 years) from one football team (f=28, m=37) and one ice hockey team (f=21, m=29) were assessed for self-reported symptoms and severity of symptoms, cognitive testing for 10-word recall and digit backwards. Oculomotor performance was assessed using eye-pursuits. Neurophysiology was assessed using transcranial magnetic stimulation. Baseline data was tested during pre-season for all athletes and, when a concussion was identified, carried out at 48h, 12-, 19-, and 26-days post injury.

**Results:** Twenty-two players suffered concussion injuries (f=10, m=12). Significant increases in symptoms were reported at 48-hours (*p*<0.001) and 12-days (*p*=0.017). Severity of symptoms were significantly increased at 48-hours only (*p*=0.002). Significant decrements in performance for 10-word recall (p=0.003), digit backwards (p=0.011), and eye-pursuit (p=0.009) were observed at 48-hours in comparison to baseline. Transcranial magnetic stimulation revealed significantly increased cortical inhibition at 48-hours (*p*=0.024), 12- (*p*=0.009), and 19-days (*p*=0.028) compared to baseline. No differences were seen between females and males for any variables or timepoints.

**Conclusion:** Concussed players show acute changes in cortical inhibition, resolving by 26-days after injury, which follows a longer time-course for recovery compared to symptoms, cognitive tests, and oculomotor eye-pursuits. These results suggest that measuring the recovery of concussed athletes should incorporate a range of testing modalities in the determination of a player’s readiness to return to play.

## Introduction

Sports-related concussion (SRC) is a common injury that occurs across a wide variety of contact team sports and affects athletes from professional to recreational levels of participation. Well acknowledged in the historical medical literature,^1^ the last 15 years in particular has seen a notable increase in concern with concussion internationally, particularly with attribution towards risk of cognitive impairments and neurodegenerative diseases.^2, 3^ While focus has been on the ‘diagnostics’ of SRC, insofar as whether concussed athletes are being appropriately identified and assessed on the day of play, increasing attention is now being directed towards recovery time-periods after SRC and the return-to-sport strategy.^4^

Clinical recovery following sports concussion has traditionally been assessed via symptom observation (number and severity), cognitive testing (involving orientation, concentration, and memory) and gross motor assessments (such as balance).^4^ Oculomotor testing has been recently suggested as part of a multi-modality clinical approach for quantifying SRC recovery.^4, 5^ Conversely, little research has investigated the physiological responses and recovery after SRC. While concussion induces a neurometabolic cascade^6, 7^, observed through functional impairments, how this affects neurophysiological post-synaptic excitability and inhibition still requires investigation. Moreover, questions remain of these effects on neuromuscular pathways, particularly when athletes report being asymptomatic.^6, 7^ This is an important line of inquiry as systematic reviews of epidemiological studies have shown significant increased risks of further injury following SRC.^8–10^

Using a neurophysiological approach can provide insight into the neuromuscular system. Transcranial magnetic stimulation (TMS) studies provide a non-invasive method to investigate the integrity of the human central nervous system via motor evoked potentials (MEPs) in SRC studies.^11–13^ A meta-analysis of TMS studies and SRC^13^ has shown that there is an increase in cortical inhibition in the acute period following injury (up to two weeks), which continues for up to 12 weeks. However, limitations of this meta-analysis include the cross-sectional design of the studies, rather than repeated measures, and the high heterogeneity of the sample.^13^ The limited number of TMS studies using a repeated-measures design appear to show a dynamic response in cortical inhibition. For example Di Virgillio et al^11^ reported increased cortical inhibition immediately following a bout of soccer heading (not concussed), returning to baseline by 24 hours. Preliminary work by Pearce et al^12^ showed in Australian football players with SRC that cortical inhibition remained increased 10 days following injury.

While TMS has the potential to be an appropriate method to gain insight into the acute pathophysiological processes of concussion, further studies are required to quantify the time-course dynamic changes using repeated-measures designs. The aim of this study was to extend on the preliminary study by Pearce et al^12^ by measuring symptomology, oculomotor eye-pursuits, and TMS in both male and female athletes up to four weeks post-SRC using a repeated-measures design.

## Methods

### Study cohort

Female and male athletes (n=115; mean age 26.2 ± 5.4 years) across two sports, Australian football and ice hockey (**Table 1**), were recruited for their respective 2023 seasons. Of the 115 participants screened, one player was excluded from baseline testing due to not meeting the inclusion criteria. Participants were from one football club participating within their sub-elite regional division, and one ice hockey club that participates in the Australian Ice Hockey League. Based on published concussion incidence rates across both sports^14–16^ we calculated an injury sample size of n=21. Written informed consent from individual participants and clubs were obtained prior to data collection. All methods were approved by the University Human Research Ethics Committee (HREC21342). Players were not involved in the design of the research or conduct of the research but were involved in the dissemination plans. Players themselves were provided individual feedback reports when requested (but were not given feedback reports of other athletes).

**Table 1.** Participants (by sports) descriptive data. Data is presented as mean (±SD).

| **Sport** | **Sex**<br>**(n)** | **Age (years)** | **Previous career**<br>**concussion history** |
| --- | --- | --- | --- |
| Australian football<br>(n=64) | F=27 | F=28.0 (6.00) | F= 9/28; 1.3 (0.7) |
|  | M=37 | M=23.90 (3.80) | M= 6/37; 1.8 (0.5) |
| Ice Hockey (n=50) | F=21 | F=28.50 (6.10) | F= 8/21; 2.5 (1.1) |
|  | M=29 | M=26.20 (4.30) | M= 5/29; 2.4 (1.9) |

Participants completed pre-screening for prior concussion history and suitability for TMS. Any individual who had a concussion within the previous six months or were currently taking prescribed medications for a neurological or psychiatric condition were excluded from the study. Baseline study protocols were completed during the preseason periods for each sport and were completed in one 45-minute visit (including pre-screening and concussion history, basic cognitive and oculomotor testing, and TMS). Players diagnosed with a SRC by their team clinician were brought in for a further four visits at 48-hours, 12-, 19-, and 26- days after the concussion. At the time of the data collection, players across both sports were not obliged to complete a 21-day rest period as recommended by the Australian Sports Commission^17^ and therefore some players were medically cleared to return to play and did not complete all data points. Athletes that were not concussed (n=52) returned to be tested within the following two-weeks after the completion of their season.

### Outcome measurements

Symptom and cognitive testing were completed using the Sport Concussion Assessment Tool V5.^18^ Symptoms were recorded as number of self-reported symptoms (maximum 22 symptoms) and severity of each identified symptom (maximum 132 points). Working memory was assessed using the 10-word recall involving three repeat trials and was scored as the number of correct responses from each trial of 10 words for three attempts (maximum score 30). Concentration was measured using the digit backwards assessment where participants were instructed to respond to a set of numbers, from three digits to a maximum of six digits, in the opposite order presented to them. Scoring for concentration included both trials (for each level of difficulty), thereby measuring scores out of eight, rather than four.

Oculomotor function was evaluated via the EyeGuide Focus (Lubbock, TX, USA) that measures eye-pursuits. Following EyeGuide recommended protocols, participants sat with their head positioned in a chin/head rest while tracking a moving white dot against a black background in an anticlockwise, then clockwise direction in a lemniscate path for 10 s. The fixed-position digital camera follows the participant’s pupils at a 60-Hz frequency, comparing the position and movement speed of the pupils to the white dot stimulus.^19^ The EyeGuide Focus ‘score’ was calculated as the cumulative distance between the stimulus data and the pupil position.^20^ Lower scores, presented in arbitrary units (AUs), were indicative of better overall performance.^21^

Neurophysiology assessment was completed using single-pulse TMS.^22^ Using well established protocols,^12, 23, 24^ TMS was applied over the contralateral motor cortex area projecting to the participants first dorsal interosseous (FDI) muscle. Surface electrodes (ADInstruments, Australia) with an inter-electrode distance of 2 cm were placed over the FDI of the participant’s dominant hand following the recommendations of the surface electromyography for non-invasive assessment of muscles (SENIAM) project.^25^ Signals were amplified (×1,000), filtered (10–1,000 Hz), and sampled at 2 kHz,^25^ recording 500 ms responses (100 ms pre-stimulus, 400 ms post-stimulus; PowerLab 4/35, ADInstruments, Australia). All TMS procedures adhered to the TMS checklist for methodological quality.^26^

A MagPro Compact stimulator (MagVenture, Denmark) with a C-B60 butterfly coil (outer dimension 75 mm) was used to generate MEPs. During TMS testing, participants wore a fitted cap (EasyCap, Germany), positioned with reference to the nasion-inion and interaural surface anatomical landmarks. The cap was marked with sites at 1-cm spacing in a latitude-longitude configuration to ensure coil position reliability for repeated testing.^12, 23, 24^ Following identification of the participant’s ‘optimal site’, where the largest MEP was observed, active motor threshold (aMT) was ascertained during a controlled, low-level voluntary contraction (10 ± 3% maximal voluntary contraction) of the FDI muscle. Stimulator output was increased in 5% steps, starting from 15% of the stimulator output, while the participant held the isometric contraction until discernible MEPs were observed.^11, 27^ Once aMT was identified, 20 stimuli (four sets of five stimuli) were delivered at 130% of aMT. To avoid stimulus anticipation, stimuli were spaced 4-8 s apart with 30-s rest between each set to reduce muscle fatigue. The same optimal site was used at subsequent visits if the participant was concussed.

Corticospinal latency was measured from the TMS stimulus artefact to the onset of the MEP waveform.^12^ MEP waveform amplitude, reflecting excitability, was calculated from the peak-to-trough difference. Duration of the cSP, indicating inhibition, was quantified from the initial onset of the MEP waveform to the return of uninterrupted EMG.^12, 23^

It has been argued that the most influential confounding factor on cSP duration is the preceding MEP. Therefore, to reflect the balance between inhibitory and excitatory mechanisms observed in the MEP waveform, cSP:MEP ratios were used to compare concussed individuals across repeated measures, and to reduce between-participant variability.^28, 29^ SRC cohorts using cSP:MEP ratios have been previously published.^30–32^

### Data and statistical analysis

Data was screened for normality using Shapiro-Wilk tests and found to have normal distributions (*W*=0.910 – 0.945; *p*>0.05). For athletes who received a concussion during the season, mixed-design ANOVAs were used to test for interactions and main effects of sex (female, male) and time (baseline, 48 h, 12-days, 19-days, 26-days). Where significant interactions or main effects were found, *post-hoc* pairwise comparisons were performed with Bonferroni corrections. In the non-concussed control group, paired *t* tests were used to assess data pre- and post-season. As only pre- and post-season data was collected in non-concussed players, this data was not directly compared to the concussed group. Alpha was set at 0.05. Data are presented as mean (±SD).

## Results

During the course of their respective seasons, 22 players were diagnosed with SRC by their team clinician (Australian football: 9 females, 7 males; ice hockey: 1 female, 5 males). All participants completed testing sessions with no adverse effects.

Self-reported symptoms (number and severity scores), 10-word recall, and reverse digits are presented in **Table 2**. There were no interactions nor main effects of sex for any variable (p>0.05). A main effect for time was observed for number of symptoms (*F*_4,44_=31.81; *p*<0.001) with the number of reported symptoms at 48-hours (p<0.01) and at 12-days (p=0.007) significantly elevated compared to baseline. Similarly, there was a main effect of time (*F*_4,44_=19.93; *p*<0.001) for symptom severity, with symptom severity scores being significantly increased at 48-hours *(t*=-5.63; *p*=0.002), compared to baseline. A main effect of time was also observed for 10-word recall (*F*_4,44_=14.21; *p*<0.001) and digit backwards (*F*_4,44_=9.08; *p*<0.001) with *post-hoc* comparisons showing significant decrement for 10-word recall and digit backwards measures at 48-hours compared to baseline (p=0.001 and p=0.039, respectively). Non-concussed players showed no differences in symptom scores, 10-word recall, or digit backward performance between pre- and post-season (all *p*>0.05; **Table 2**).

**Table 2.**
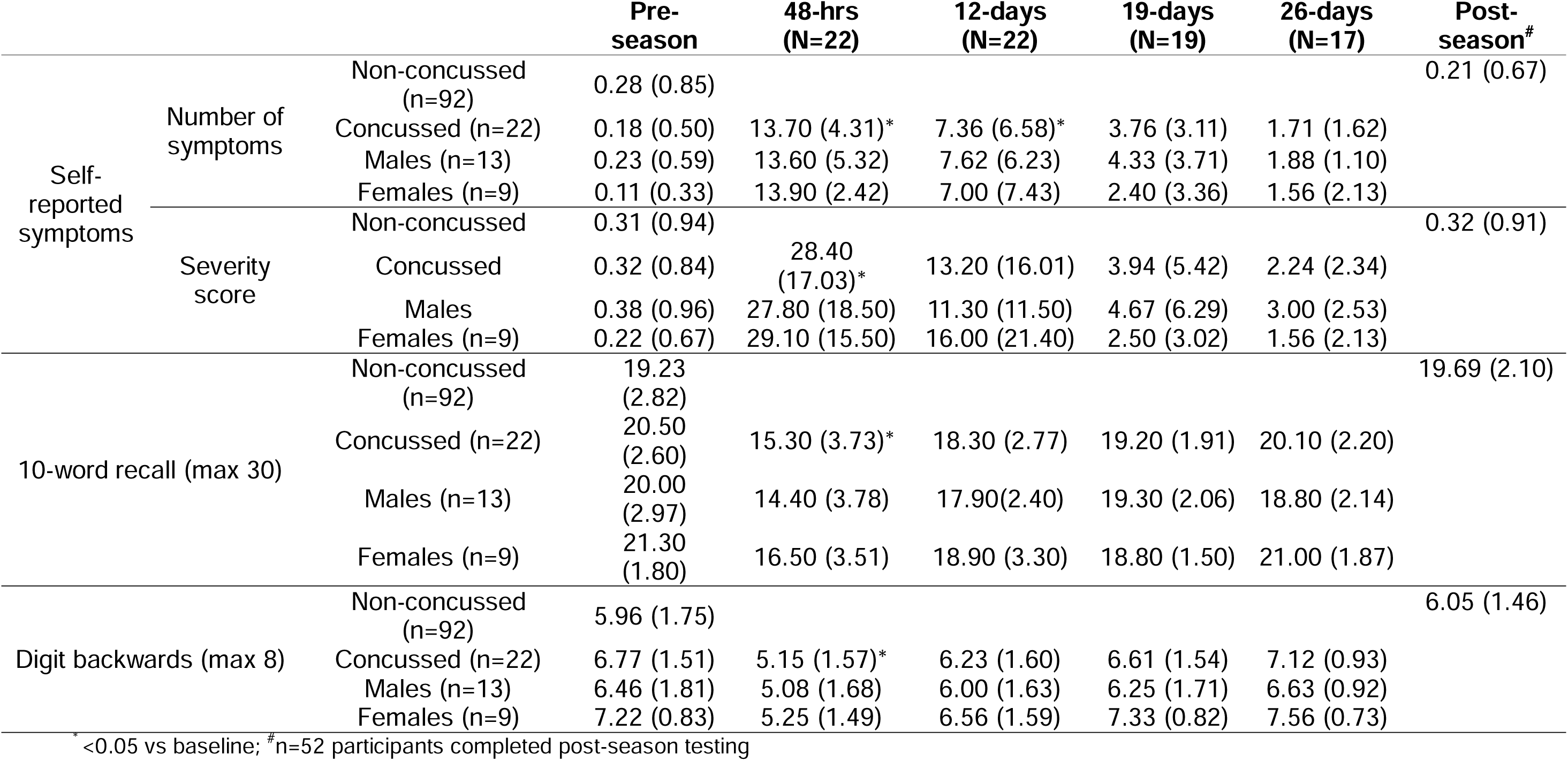
Self-reported symptoms, 10-word recall, and digit backward results by overall group and sex. Mean (±SD)

Oculomotor data are illustrated in **Figure 1**. There was no interaction (*p*=0.554) nor main effect of sex (*p*=0.962). A main effect for time was observed (*F*_4,44_=11.11; *p*<0.001) with *post-hoc* analyses showing a significantly higher score (greater error) at 48-hours compared to baseline (*p*=0.009). For comparison, non-concussed players’ data for pre- and post-season is also presented in **Figure 1**. No changes were observed in non-concussed players (*p*=0.729).

**Figure 1.**
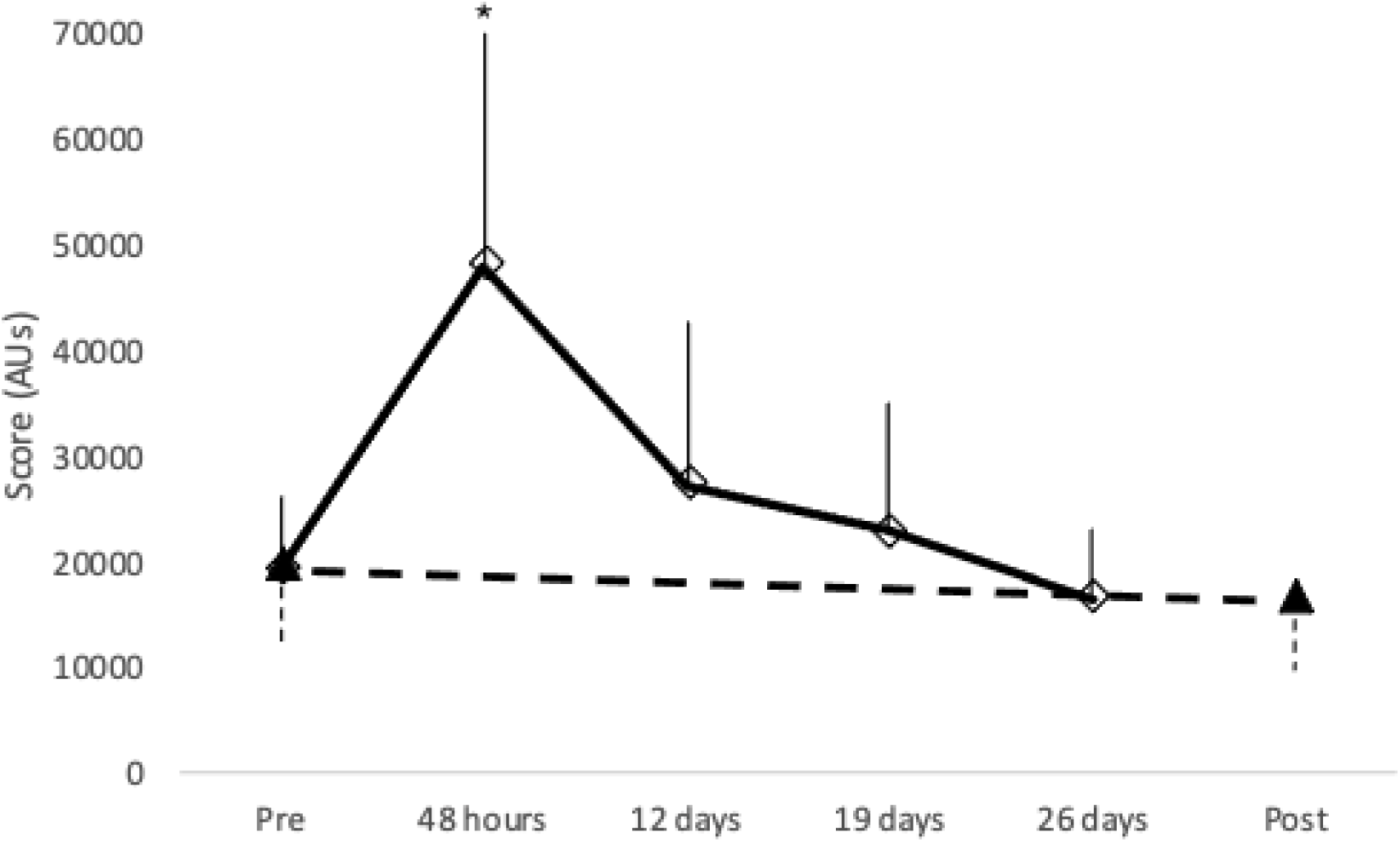
EyeGuide Focus score (mean ±SD) between concussed (open diamond, solid line) and non-concussed control (closed triangle, dashed line) groups. AUs=Arbitrary Units. *p<0.05 vs pre.

Evoked potential TMS data is presented in **Figure 2**. There was no interaction (*p*=0.372) nor main effect of sex (*p*=0.610). A main effect for time was found (*F*_4,44_=10.92; *p*<0.001) with *post-hoc* comparisons showing a significant increase in cSP:MEP ratio, suggesting increased cortical inhibition, at 48-hours (*p*=0.024), 12- (*p*=0.009) and 19-days (*p*=0.024) compared to baseline. **Figure 3** provides an illustrative example of MEP sweeps from one participant. There were no interactions nor main effects for aMT or latency (p>0.05). Non-concussed players’ cSP:MEP ratio did not show differences pre- to post-season (p=0.352).

**Figure 2.**
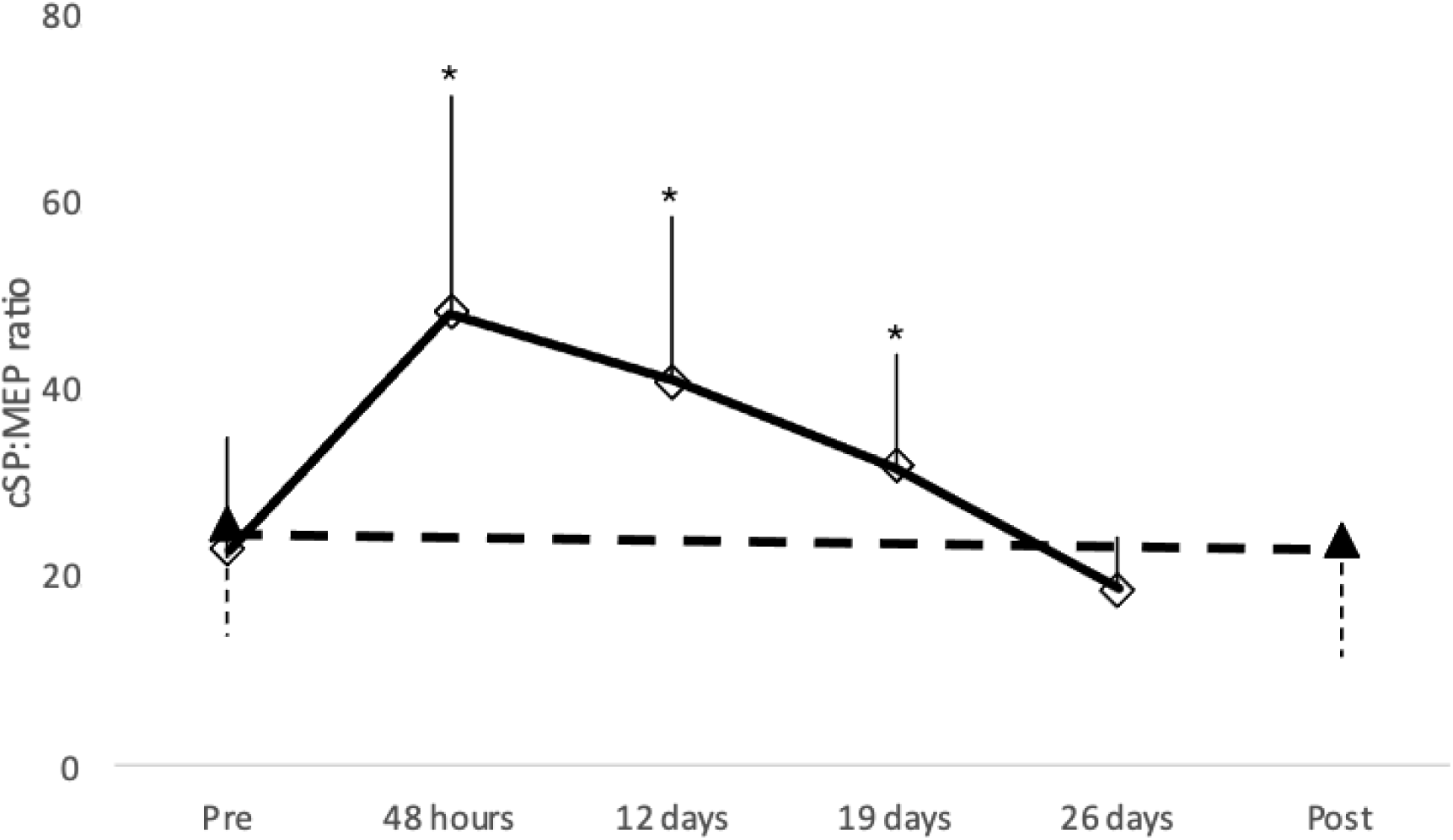
TMS cSP:MEP ratio (mean ±SD) between (a) concussed (open diamond, solid line) and non-concussed (closed triangle, dashed line) control groups. *p<0.05 vs pre.

**Figure 3.**
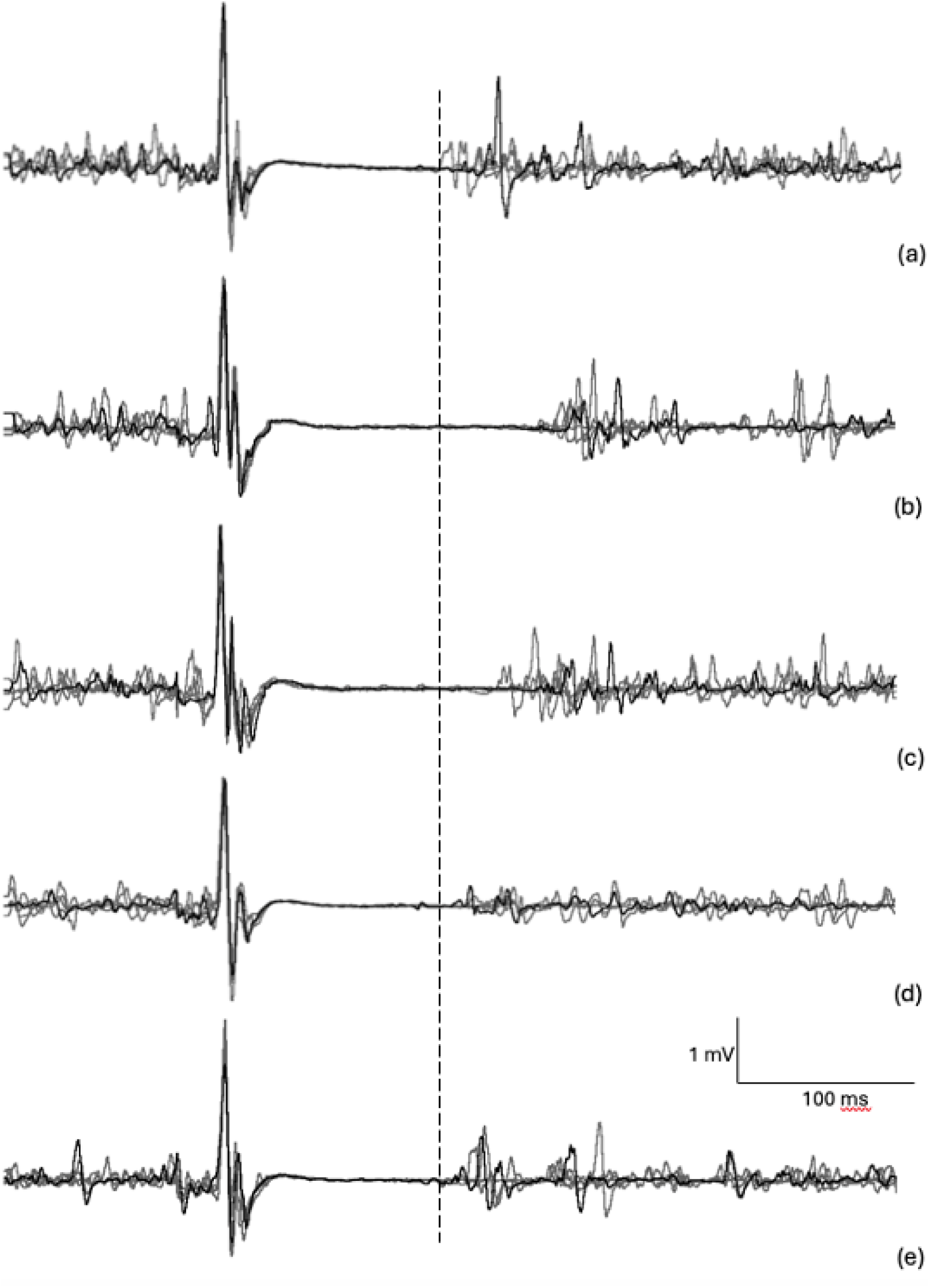
Example of overlaid MEP sweeps (500 ms) showing increased inhibition via longer silent period duration. Overlaid MEP sweeps illustrate pre-season baseline (a), and post-concussion at 48-hours (b), 12-days (c), 19-days (d), and 26 days (e). Dashed line indicates comparison to baseline.

## Discussion

The aim of this study was to investigate the time-course recovery of symptoms, cognitive testing, oculomotor, and neurophysiological responses following concussion in both male and female athletes. Our data showed that while symptoms, cognitive, and oculomotor performance resolved within 12-days, neurophysiological measures returned to baseline by 26-days. We also found no effect of sex on the time-course for recovery of these variables. As such, return-to-play protocols following concussion injury should not rely only on symptom reporting or oculomotor function, as full recovery likely has not been obtained and may require longer than the typical periods often used in concussion guidelines.^4^

Assessment for concussion recovery typically utilise symptom reporting and cognitive and gross motor testing to assess recovery following concussion, with the most recent consensus statement also supporting the use of oculomotor testing.^4^ Oculomotor testing may be advantageous to traditional symptom reporting and performance in memory and concentration tests as it provides objective data that is harder for athletes to manipulate, which Australian athletes have previously admitted.^33, 34^ While these concussion assessment tools generally are able to show immediate deficits in cognitive or eye-pursuit performance, it has been reported that sensitivity diminishes as time progresses.^5^ For example, Patricious et al^4^ noted that the psychometric utility of the SCAT weakens after 72 hours to which an office-based assessment tool has since been developed to account for this.^35^ Similarly, in the case of EyeGuide Focus, while *intra*-session testing has shown moderate to good reliability,^19^ poor reliability has been reported with *inter*-session testing.^21^ Using a different oculomotor test that uses saccades (rather than smooth pursuit), such as the King-Devick test, shows good *inter*-session reliability^36^ and reports oculomotor deficits following concussion in female rugby players up to 28 days post injury.^37^ Future studies using oculomotor testing could benefit from incorporating both saccades and eye-pursuits within a multimodality testing battery for concussion.

The finding of increased cortical inhibition following concussion is consistent with previous research^13^ may reflect a possible number of mechanisms. For example, Giza and Hovda^6, 7^ suggest concussion induces a mechanical stretching of axons that may in turn contribute to membrane disruption, affecting neural transmission that can persist for weeks. They also hypothesise that concussion is the result of a cortical neurometabolic cascade, affecting molecular and ionic processes, as well as alterations in neurotransmitter activity including excitatory transmitters such as glutamate, but also the inhibitory neurotransmitter GABA.^6, 7^ This neurometabolic cascade consequently affects neural responses that can be seen in the MEP waveform, specifically a lengthened cSP (see example **Figure 3**) indicating increased cortical inhibition.^13^ However, contribution to altered MEP waveforms may also originate from inflammation in glial cells impacting neuronal physiology.^38^

Within the broader discussion on the potential advantages of objective measures in concussion, MEPs provide a unique opportunity to quantify neurophysiological changes that may negatively impact on descending motor pathways. Data from this study, concurring with previous TMS research^12, 39^ showing imbalances towards increased cortical inhibition, could provide a reason for the increased risk of musculoskeletal injuries following concussion.^9, 10, 40^ While not specifically measuring variables of motor control changes in this study, previous TMS concussion studies have demonstrated slowed reaction times^12^ and cognitive processing speed^12, 39^ associated with increased inhibition. Increased inhibition may impact not only on quick decisions required during competition and training, but also neuromechanical delay. For example, the majority of anterior cruciate ligament injuries are non-contact and happen between 40-60 ms after foot contact,^41, 42^ which may contribute to the loss of neuromuscular control and coordination errors leading to increased risk of ACL injury.^10^ Similarly, previous studies using movement-related cortical potentials have shown persistent reduction in amplitudes (suggesting inhibition) up to 30-days post-concussion that contribute to postural control during movements.^43^ Collectively, these studies show that post-concussion management and return-to-sport decisions should not only include symptom observation and cognitive testing, but also neurophysiological assessment for sub-clinical time-course changes.

A secondary question of this study addressed differences in recovery times between males and females. While emerging data suggests that females self-report worse symptoms, and longer recovery times,^44, 45^ research in college athletes appears to show no differences between males and females in reporting of symptoms^46^ and gait-related motor control,^47^ results that support those of the current study. There is evidence suggesting MEP cortical inhibition is influenced by the natural menstrual cycle^48^ but blunted with contraceptive pill use^49^. Therefore, future TMS studies comparing time-course changes post-concussion between sexes may benefit from the inclusion of menstrual cycle data from female athletes.

### Limitations

At the time of the data collection, study participants were not obliged to complete a 21-day rest period as recommended by the Australian Sports Commission^17^ (February 2024) but rather utilised the Australian Football League 12-day rest period. Therefore, some players were medically cleared to return to competition and did not complete all data points. However, with the assistance of the clubs and athlete participation, compliance for measures at the 19- and 26-day timepoints were 86% and 77% respectively.

### Research Implications

These results emphasise the importance of including physiological measures, such as evoked potentials, further to symptomology and cognitive measures, in progressing concussion research investigating return to play timelines. Further, the implications of the research also contribute to informing continuing public discourse on safe return to sport timelines following concussion, and the importance of athletes not being hurried to return to competition given the increased risk of further injury that may be due to transient neurophysiological impairments as reported in this study.

## Conclusion

In conclusion, this multi-modality study incorporating neurophysiological measures from TMS in addition to more established concussion testing protocols, has shown changes in cortical inhibition persisting beyond that of clinical symptoms and oculomotor assessment. Data in this study not only concurs with previous TMS studies^12, 39^ but also recent blood biomarker research illustrating neurophysiological recovery taking a longer time-course than that of clinical recovery.^50^ Consequently, having additional rigour in the testing of athletes will address concerns regarding successful hiding of symptoms by athletes^33, 34^ as well as uncertainty on symptom self-report and observation in return-to-sport decisions.

## Acknowledgements

The authors would like to thank Mr Dale O’Neil, Ms Emily Verdouw, Mr Andrew Erzen, and Ms Keira Dunwoody for assistance with organisation of pre-season and follow-up testing.

## Contributors

All authors contributed to the creation, study proposal, design, data analysis, and manuscript drafts. AJP led the data collection. AC contributed to data collection. All authors have read the final manuscript and approved the manuscript to be published.

## Ethics approval

La Trobe University granted ethical approval for this study (HREC21342). This study was conducted with the ethical standards of the Helsinki Declaration (1964) and later amendments. Data management was consistent with La Trobe University’s data management policy.

## Data availability statement

Data are available from the corresponding author upon reasonable request and with relevant University Human Research Ethics approval.

## Biographical note

AJP is a neurophysiologist with a research focus on the neurophysiology of concussion and chronic traumatic encephalopathy. KM is a biomechanist with a research focus on physical performance and biomechanical modelling of human movement. AC is a sports physiologist focusing research on performance, health, and wellbeing of female athletes.

